# Evaluation of Nucleocapsid and Spike Protein-based ELISAs for detecting antibodies against SARS-CoV-2

**DOI:** 10.1101/2020.03.16.20035014

**Authors:** Wanbing Liu, Lei Liu, Guomei Kou, Yaqiong Zheng, Yinjuan Ding, Wenxu Ni, Qiongshu Wang, Li Tan, Wanlei Wu, Shi Tang, Zhou Xiong, Shangen Zheng

**Affiliations:** Department of Transfusion, General Hospital of Central Theater Command of the People’s Liberation Army, Wuhan, Hubei, China; Department of Disease Control and Prevention, General Hospital of Central Theater Command of the People’s Liberation Army, Wuhan, Hubei, China

**Keywords:** COVID-19 diagnosis, ELISA, antibody, IgM, IgG, nucleocapsid protein, spike protein

## Abstract

**Background:** At present, PCR-based nucleic acid detection cannot meet the demands for coronavirus infectious disease (COVID-19) diagnosis.

**Methods:** 214 confirmed COVID-19 patients who were hospitalized in the General Hospital of Central Theater Command of the People’s Liberation Army between January 18 and February 26, 2020, were recruited. Two Enzyme-Linked Immunosorbent Assay (ELISA) kits based on recombinant SARS-CoV-2 nucleocapsid protein (rN) and spike protein (rS) were used for detecting IgM and IgG antibodies, and their diagnostic feasibility was evaluated.

**Results:** Among the 214 patients, 146 (68.2%) and 150 (70.1%) were successfully diagnosed with the rN-based IgM and IgG ELISAs, respectively; 165 (77.1%) and 159 (74.3%) were successfully diagnosed with the rS-based IgM and IgG ELISAs, respectively. The positive rates of the rN-based and rS-based ELISAs for antibody (IgM and/or IgG) detection were 80.4% and 82.2%, respectively. The sensitivity of the rS-based ELISA for IgM detection was significantly higher than that of the rN-based ELISA. We observed an increase in the positive rate for IgM and IgG with an increasing number of days post-disease onset (d.p.o.), but the positive rate of IgM dropped after 35 d.p.o. The positive rate of rN-based and rS-based IgM and IgG ELISAs was less than 60% during the early stage of the illness 0-10 d.p.o., and that of IgM and IgG was obviously increased after 10 d.p.o.

**Conclusions:** ELISA has a high sensitivity, especially for the detection of serum samples from patients after 10 d.p.o, it can be an important supplementary method for COVID-19 diagnosis.

## Introduction

The ongoing outbreak of coronavirus infectious disease 2019 (COVID-19) (1), which emerged in Wuhan, China, is caused by a novel coronavirus named severe acute respiratory syndrome coronavirus 2 (SARS-CoV-2) (2-4). As of March 1, 2020, more than 80,000 laboratory-confirmed cases have been reported in China (5), and the disease has spread over 58 countries in Asia, Australia, Europe, and North America (1). On January 30, 2020, the WHO declared the outbreak of COVID-19 a Public Health Emergency of International Concern (PHEIC).

SARS-CoV-2 is the seventh member of enveloped, positive-stranded RNA viruses (4) that are able to infect humans. Genomic characterization of SARS-CoV-2 identified it as a Beta coronavirus and showed it is closely related (with 96% identity) to Bat CoV RaTG13, but distinct from SARS-CoV (6). SARS-CoV-2 has a receptor-binding domain (RBD) structure similar to that of SARS-CoV. Functionally important ORFs (ORF1a and ORF1b) and major structural proteins, including the spick (S), membrane (M), envelope (E), and nucleocapsid (N) proteins, are also well annotated (7). According to previous reports, the M and E proteins are necessary for virus assembly (8, 9). The S protein is important for attachment to host cells, where the RBD of S protein mediates the interaction with angiotensin-converting enzyme 2 (ACE2) (6). The S protein is located on the surface of the viral particles and has been reported to be highly immunogenic (10). The N protein is one of the major structural proteins of the virus and is involved in the transcription and replication of viral RNA, packaging of the encapsidated genome into virions (11, 12), and interference with cell cycle processes of host cells (13). Moreover, in many coronaviruses, including SARS-CoV, the N protein has high immunogenic activity and is abundantly expressed during infection (14-16). Both S and N proteins may be potential antigens for serodiagnosis of COVID-19, just as many diagnostic methods have been developed for diagnosing SARS based on S and/or N proteins (10, 14-17).

Currently, diagnosis of COVID-19 is confirmed by RNA tests with real-time (RT)-PCR or next-generation sequencing. Studies have shown that SARS-CoV-2 mainly infects the lower respiratory tract, and that viral RNA can be detected from nasal and pharyngeal swabs and bronchoalveolar lavage (BAL) (3, 6, 18). However, the collection of the lower respiratory samples (especially BAL) requires both a suction device and a skilled operator. A previous study showed that except for BAL, the sputum from confirmed patients possessed the highest positive rate, ranging from 74.4% to 88.9%. The positive rate of nasal swabs ranged from 53.6% to 73.3%, and throat swabs collected ≥8 days post-disease onset (d.p.o.) had a low positive rate, especially in samples from mild cases (19). The viral colonization of the lower respiratory tract and the collection of different respiratory specimens cause a high false negative rate of real time Reverse Transcription-Polymerase Chain Reaction (RT-PCR) diagnosis. Therefore, with the current tests it is difficult to achieve a meaningful assessment of the proportion of symptomatic patients that are infected, and a rapid and accurate detection method of COVID-19 is urgently needed. Serological assays are accurate and efficient methods for the screening for many pathogens, as specific IgM and IgG antibodies can be detected with ELISA, which has relatively high throughput capacity and less stringent specimen requirements (uniformly serum collection) than RNA-based assays.

The present study was conducted to evaluate the performance of rN-based and rS-based ELISAs to detect IgM and IgG antibodies in human serum against SARS-CoV-2. A total of 214 clinical serum samples from confirmed COVID-19 patients and 100 samples from healthy blood donors were tested with the rN-based and rS-based ELISAs.

## Materials and Methods

### Patients and samples

A total of 214 patients diagnosed with COVID-19, hospitalized in the General Hospital of the Central Theater Command of the People’s Liberation Army (PLA) between January 18 and February 26, 2020, were recruited. The general information was extracted from electronic medical records. All of the patients were laboratory-confirmed positive for SARS-CoV-2 by RT-PCR using pharyngeal swab specimens, and at a median of 15 d.p.o. (range, 0–55 days). Serum samples from 100 healthy blood donors were selected as controls. This study was approved by the Hospital Ethics Committee of the General Hospital of the Central Theater Command of the PLA ([2020]003-1) and the written informed consent was waived for emerging infectious diseases.

### rN-based ELISA

The recombinant nucleocapsid (rN) protein-based ELISA kit (Lizhu, Zhuhai, China) was used for the detection of IgM or IgG antibody against SARS-CoV-2. For IgM detection, ELISA plates were coated with monoclonal mouse anti-human IgM (μ chain) antibody. Serum sample (100 μL, diluted 1:100) was added to the pre-coated plates, and plates were incubated at 37 °C for 1 h. Three replicates of each sample or control were included on each plate. After washing, 100 μL horseradish peroxidase (HRP)-conjugated rN protein of SARS-CoV-2 was added. Then, the plate was incubated at 37 °C for 30 min, followed by washing. TMB substrate solution (50 μL) and the corresponding buffer (50 μL) were added, and samples were incubated at 37 °C for 15 min. The reaction was terminated by adding 50 μL of 2 M sulfuric acid, and the absorbance value at 450 nm (A_450_) was determined. The cutoff value was calculated by summing 0.100 and the average A_450_ of negative control replicates. When A_450_ was below the cutoff value, the test was considered negative, and when A_450_ was greater than or equal to the cutoff value, the test was considered positive.

For IgG detection, ELISA plates were coated with rN protein. Serum sample (5 μL) diluted in 100 μL dilution buffer was added to the plates. After incubation and washing, HRP-conjugated monoclonal mouse anti-human IgG antibody was added to the plates for detection. The other operation steps were performed as described above for IgM detection. The cutoff value was calculated by summing 0.130 and the average A_450_ of negative control replicates. When A_450_ was below the cutoff value, the test was considered negative, and when A_450_ was greater than or equal to the cutoff value, the test was considered positive.

### rS-based ELISA

The rS-based ELISA kit (Hotgen, Beijing, China) was developed based on the RBD of the recombinant S polypeptide (rS). For IgM antibody testing, serum samples (diluted 1:100) and negative and positive controls were added to the wells of the rS-coated plates in a total volume of 100 μL, and plates were incubated at 37 °C for 30 min. After five wash steps with washing buffer, 100 μL of diluted HRP-conjugated anti-human IgM antibodies was added to the wells, and samples were incubated at 37 °C for 30 min. After five wash steps with washing buffer, 50 μL of TMB substrate solution and 50 μL of the corresponding buffer were added, and samples were incubated at 37 °C for 10 min. The reaction was terminated by adding 50 μL of 2 M sulfuric acid, and A_450_ was measured. The IgG antibody was determined by a standard ELISA procedure as described above, except that serum sample was diluted 1:20, and the detector was HRP-conjugated anti-human IgG. The cutoff values of IgM and IgG were calculated by summing 0.250 and the average A_450_ of negative control replicates. When A_450_ was below the cutoff value, the test was considered negative, and when A_450_ was greater than or equal to the cutoff value, the test was considered positive.

### Statistical analysis

Categorical variables were expressed as the counts and percentages and compared using the chi-square test, while the Fisher exact test was used when the data were limited. Statistical analyses were performed using SPSS version 22.0. A two-sided *P*-value < 0.05 was considered statistically significant.

## Results

### Performance of serological assays by rN-based and rS-based ELISA

The serum samples were collected from 214 COVID-19 patients who were confirmed by qRT-PCR and hospitalized in the General Hospital of the Central Theater Command of the PLA. Of the 214 serum samples, 146 (68.2%) and 150 (70.1%) were identified as positive by the rN-based IgM and IgG ELISAs (Table 1), respectively. IgM and/or IgG means that one of them or both were detected in serum samples; 172 (80.4%) serum samples were positive by rN-based ELISA (Fig. 1). The numbers of positive results with IgM, IgG, and IgM and/or IgG detected by rS-based ELISA were 165 (77.1%), 159 (74.3%), and 176 (82.2%), respectively (Table 1 and Fig. 1).

**Table 1.** Positive rate of rN-based and rS-based ELISA for detection of IgM and IgG in serum samples of patients at different stages after disease onset.

| Days | No. of serum samples | No. (%) positive for IgM |  |  | No. (%) positive for IgG |  |  | No. (%) positive for IgM and/or IgG |  |  |
| --- | --- | --- | --- | --- | --- | --- | --- | --- | --- | --- |
|  |  | rN-based | rS-based | P | rN-based | rS-based | P | rN-based | rS-based | P |
| Total | 214 | 146(68.2) | 165(77.1) | 0.039 | 150(70.1) | 159(74.3) | 0.332 | 172(80.4) | 176(82.2) | 0.620 |
| 0-5 | 22 | 7(31.8) | 8(36.4) | 0.750 | 7(31.8) | 9(40.9) | 0.531 | 9(40.9) | 10(45.5) | 0.761 |
| 6-10 | 38 | 20(52.6) | 19(50.0) | 0.818 | 15(39.5) | 19(50.0) | 0.356 | 20(52.6) | 23(60.5) | 0.488 |
| 11-15 | 54 | 39(72.2) | 45(83.3) | 0.165 | 39(72.2) | 41(75.9) | 0.661 | 48(88.9) | 49(90.7) | 0.750 |
| 16-20 | 55 | 45(81.8) | 53(96.4) | 0.014 | 48(87.3) | 51(92.7) | 0.340 | 52(94.5) | 53(96.4) | 0.647 |
| 21-30 | 32 | 26(81.3) | 28(87.5) | 0.491 | 28(87.5) | 27(84.4) | 0.719 | 30(93.8) | 28(87.5) | 0.391 |
| 31-35 | 6 | 5(83.3) | 6(100.0) | 0.296 | 6(100.0) | 5(83.3) | 0.296 | 6(100.0) | 6(100.0) | N/A |
| >35 | 7 | 4(57.1) | 6(85.7) | 0.237 | 7(100.0) | 7(100.0) | N/A | 7(100.0) | 7(100.0) | N/A |

IgM and/or IgG: at least a positive result detected by IgM and IgG ELISA
Days: Days post-disease onset.
N/A: Not available.

**Figure 1.**
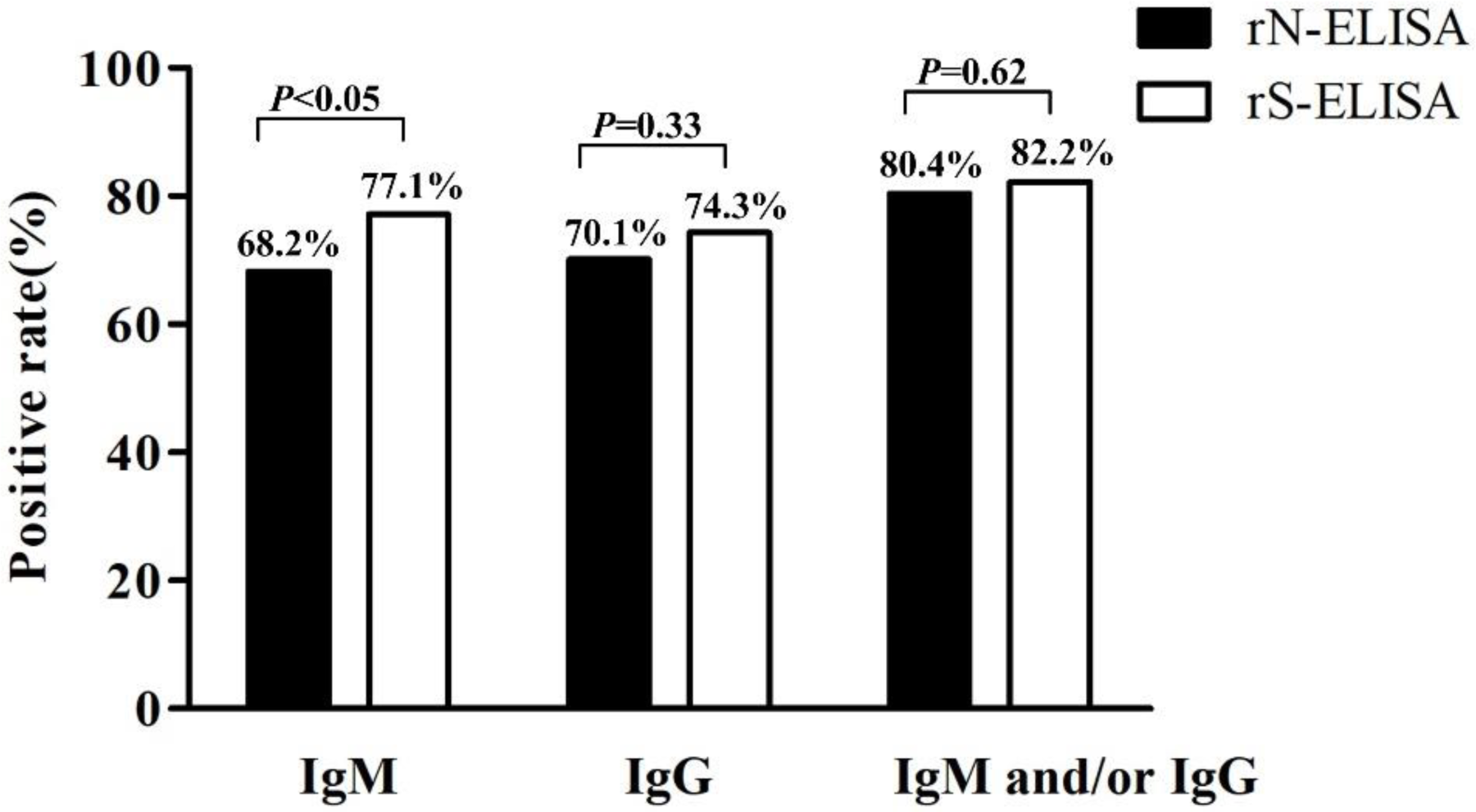
Comparison of positive rate of antibodies detected by rN-based ELISA and rS-based ELISA. IgM means a positive result of IgM antibody detection by ELISA, IgG means a positive result of IgG antibody detection by ELISA, IgM and/or IgG means at least one of them was positive by IgM and IgG ELISA. Results were compared by chi-square tests.

To study the production of antibodies in COVID-19 patients, we analyzed the positive rates of IgM and IgG in serum samples of all patients post-disease onset. Based on the number of days from disease onset to serum collection, patients were divided into seven groups: 0–5, 6–10, 11–15, 16–20, 21–30, 31–35, and >35 d.p.o. The median number of d.p.o. of serum sample collection was 15 (range, 0–55 days). The positive rates of IgM and IgG detected by the rN- and rS-based ELISAs in different groups are shown in Table 1. For rN-based ELISA, a clear increase in IgM and IgG positive rates was observed (Fig. 2a). The positive rates of IgM and IgG were low at 0–5 d.p.o. and 6–10 d.p.o., and the positive rate of IgM was higher than that of IgG at 6–10 d.p.o. and much lower after 35 d.p.o. (Fig. 2a), illustrating the dynamic pattern of acute viral infection where IgG concentrations rise as IgM levels drop. The IgM and/or IgG positive rate was 88.9% at 11–15 d.p.o., and more than 90% at later stages of the disease (Table 1).

**Figure 2.**
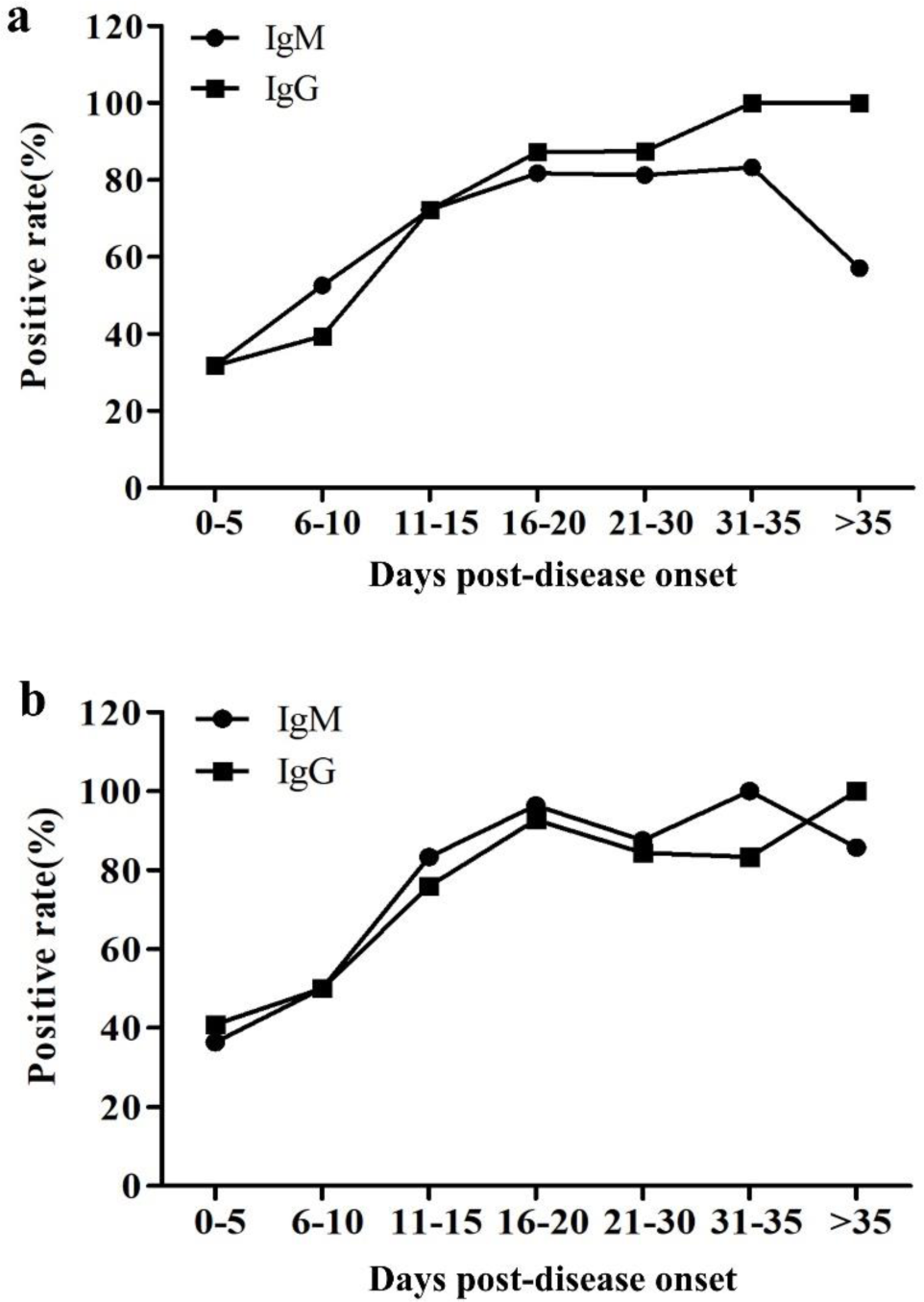
Dynamic trend of the positive rate of IgM and IgG in serum of patients at the different stage of disease. Patients were divided into seven groups of 0-5, 6-10,11-15, 16-20,21-30, 31-35 and >35 days post-disease onset, **a)** The dynamic trend of antibodies positive rate detected by rN-based ELISA. **b)** The dynamic trend of antibodies positive rate detected by rS-based ELISA.

For rS-based ELISA, a similar trend of IgM and IgG positive rates was observed (Fig. 2b). The positive rate of IgM is a little higher than that of IgG at different disease stages, except >35 d.p.o., possibly due to the higher sensitivity of rS-based IgM detection (77.1%) than IgG detection (74.3%) (Table 1).

To verify the specificity of the ELISA assays, 100 samples from healthy blood donors were analyzed. No positive result was found in both rN- and rS-based IgM and IgG ELISAs.

### Comparison of rN- and rS-based IgM and IgG ELISAs for diagnosis of SARS-CoV-2

The sensitivity of the rS-based IgM ELISA was significantly higher than that of the rN-based ELISA (*P* < 0.05). No significant difference between rS- and rN-based ELISAs was observed for detecting IgG and total antibodies (IgM and IgG) (Fig. 1). The positive rates of the rN- and rS-based ELISAs for the detection of IgM and/or IgG in serum samples collected from patients at different stages of the disease are shown in Table 1. Within 15 d.p.o., for detecting IgM, which is an important marker for early infection, rN- and rS-based ELISAs detected 57.9% (66/114) and 63.2% (72/114) of patients, indicating that the performance of the rS-based IgM immunoassay seems better than that of the rN-based assay, but there is no statistical difference between the two results. The positive rate of the rS-based ELISA for IgM is significantly higher than that of the rN-based ELISA at 16–20 d.p.o(Table 1). No significant difference between the two antigen-based ELISAs was observed for IgM detection in other groups. For IgG detection, there was no significant difference between the two kits in all groups.

In this study, the rN- and rS-based ELISAs for detecting IgM and IgG in 214 COVID-19 patients were evaluated (Table 2). In total, the detected positive rate (174/214, 81.3%) for IgM by the two kits was significantly higher than that of the rN-based ELISA kit alone (68.2%; *P* < 0.01), but was not significantly different from that of the rS-based kit alone. For IgG, 172 of 214 (80.4%) were identified positive by both kits. This sensitivity is significantly higher than that detected by only the rN-based ELISA (70.1%, *P* < 0.05) and is not significantly different from that of the rS-based ELISA.

**Table 2.**
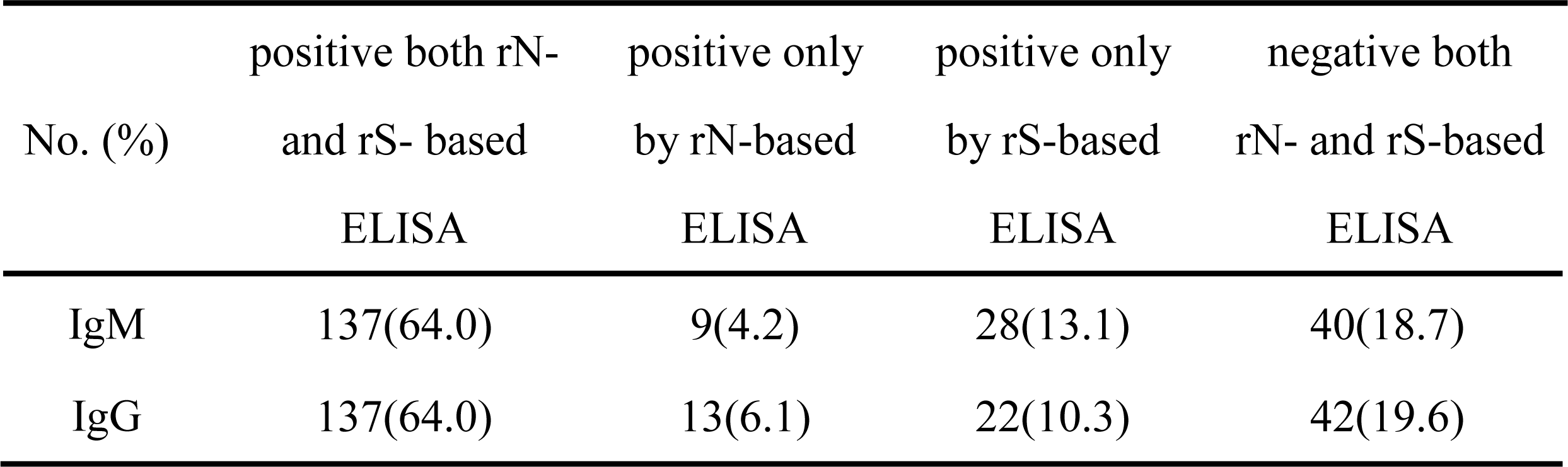
Summary results for IgM and IgG detection in the 214 serum samples from patients with COVID-19.

| No. (%) | positive both rN-and rS- based<br>ELISA | positive only<br>by rN-based<br>ELISA | positive only<br>by rS-based<br>ELISA | negative both<br>rN- and rS-based<br>ELISA |
| --- | --- | --- | --- | --- |
| IgM | 137(64.0) | 9(4.2) | 28(13.1) | 40(18.7) |
| IgG | 137(64.0) | 13(6.1) | 22(10.3) | 42(19.6) |

## Discussion

The recent outbreak and rapid spread of the novel coronavirus SARS-CoV-2 pose a great threat of a pandemic outbreak of COVID-19 to the world. Diagnostic methods are the frontline strategy for recognizing this disease. Currently, SARS-CoV-2 can be detected using RT-PCR, but inadequate access to reagents and equipment, the requirement to upscale lab facilities with restrictive bio-safety levels and technical sophistication, and the high rate of false negative results caused mainly by an unstandardized collection of respiratory specimens have resulted in low efficiency of in-time detection of the disease. Although previous studies have shown that virus-specific IgM and IgG levels allow for serologic diagnosis of SARS (15, 20), less amount of data on the serologic diagnosis using antibodies against SARS-CoV-2 are available.

In this study, we evaluated immunoassays for the detection of antibodies against SARS-CoV-2. Specifically, we evaluated IgM and IgG production and their diagnostic value. rN- and rS-based ELISAs were used to detect IgM and IgG in serum samples of confirmed COVID-19 patients. The results revealed that the rS-based ELISA is more sensitive than the rN-based one in the detection of IgM antibodies (Fig. 1). We speculate that this difference is due to the relatively high sensitivity and early response to the S antigen compared to the N antigen in patients with COVID-19. We divided the patients into different groups according to the number of d.p.o. and observed an increase in the IgM and IgG positive rate with time, except the IgM positive rate decreased at >35 d.p.o. (Fig. 2). This dynamic pattern is consistent with that observed in acute viral infection, with the IgG concentration rising gradually as IgM levels drop after one month post-disease onset. The positive rates of IgM and IgG antibodies in samples tested by the rN- and rS-based ELISAs were about 30%–50% in the 0–5 d.p.o. and 6–10 d.p.o. groups (Table 1 and Fig. 2). This may be due to the low antibody titers in early stages of the disease. Our results showed that IgM and/or IgG of SARS-CoV-2 might be positive (88.9% by rN-based ELISA, 90.7% by rS-based ELISA) at 11–15 d.p.o. (Table 1), which is to a certain extent in accordance with a previous publication about SARS, which reported that detection of antibodies to SARS-CoV could be positive as early as 8–10 d.p.o. and often occurs around day 14 (15). We observed a decreased positive rate of IgM at > 35 d.p.o.; unfortunately, we could not collect the samples from these patients after discharge. Combined, rN- and rS-based ELISAs for IgM and IgG detection were more sensitive than the rN-based ELISA alone, but there was no significant difference when compared with the rS-based ELISA. Therefore, combined use of rN- and rS-based ELISAs is not recommended for COVID-19 diagnosis; however, if an ELISA kit coated with a cocktail of N and S polypeptides shows better results, further evaluation is needed.

This study demonstrated that rN- and rS-based ELISAs can be an important screening method for COVID-19 diagnosis, with high sensitivity, especially for the analysis of serum samples from patients after more than 10 d.p.o. However, the rS-based immunoassay is recommended for early screening of suspected COVID-19 patients with negative PCR test results. The ELISA serodiagnosis can be a supplementary method to RT-PCR for COVID-19 diagnosis. ELISA can be used to quickly screen all febrile patients effectively, as large-scale confirmation or exclusion of patients is essential for controlling the disease.

## Data Availability

All data generated or analyzed during this study are included in this article

## Acknowledgments

We appreciate the helpful advice from professor Ruifu Yang from the Beijing Institute of Microbiology and Epidemiology regarding this work and his revision of this manuscript. This work was supported by the National Natural Science Foundation of China (81801984), the China Postdoctoral Science Foundation (2019M664008), and the Wuhan Young and Middle-aged Medical Backbone Talents Training Project (Wuweitong [2019] 87^th^). We thank all healthcare workers involved in this study. We thank LetPub (www.letpub.com) for its linguistic assistance during the preparation of this manuscript.

## Conflict of interest

The authors declare that no conflict of interest exists.

## Authors’ contributions

S.Z. and W.L. conceived the study and designed experimental procedures. L.L., G.K., Y.Z., Y.D., W.N., Q.W., L.T., W.W., S.T., and Z.X. collected patients’ samples. L.L. and G.K. established the ELISA and performed serological assays. W.L., S.Z., and L.L. wrote the paper. All authors contributed to data acquisition, data analysis, and/or data interpretation, and reviewed and approved the final version.

## Notes

### Competing Interest Statement

The authors have declared no competing interest.

